# ADHD Symptoms and Cannabis Use: The Role of Cannabinoid Receptor 1 and Neural Response Inhibition

**DOI:** 10.64898/2026.06.24.26356461

**Authors:** Jacqueline Aloumanis, Shanting Chen, J. Hunter Allen, Chia-Chuan Yu, Sara Jo Nixon, Amanda Elton

**Author notes:** Corresponding Author: Amanda Elton, Address: 1149 Newell Dr, Gainesville, FL 32610.

## Abstract

**Background:** Individuals with attention-deficit hyperactivity disorder (ADHD) are at increased risk for cannabis misuse, with increasing prevalence among young adults. Existing evidence suggests that cannabis can have therapeutic effects on ADHD symptoms, and continued use may be partly driven by perceived improvements in symptom-related deficits. To investigate the neural evidence for these associations, we integrated functional neuroimaging and Allen Human Brain Atlas transcriptomic data to assess neural correlates of ADHD in regions targeted by cannabinoids as predictors of cannabis use. We hypothesized that greater ADHD symptoms would lead to higher cannabis use frequency through associations of ADHD symptoms with functional deficits in cannabinoid receptor type 1 (CB1R; encoded by the *CNR1* gene) expressing brain regions.

**Methods:** We tested 466 college students (ages 18-19) with varying ADHD symptom severity and cannabis use, self-reported at baseline and three yearly-follow up questionnaires. ADHD-related neural deficits were tested in a subset of 144 participants using an fMRI stop-signal task at baseline. Growth mixture modelling categorized participants with similar cannabis use into three latent classes. The covariance between the *CNR1* gene expression map and differences in stop-signal task activation were tested as a mediator linking ADHD symptoms and cannabis use.

**Results:** Greater ADHD symptoms significantly predicted reduced activation within *CNR1*-expressing regions, which predicted higher-use cannabis class membership.

**Conclusions:** Our results add support for the self-medication hypothesis for higher rates of cannabis use among individuals with greater ADHD symptoms, which may be mechanistically linked through CB1R-enriched attention and inhibitory networks, highlighting neural targets for prevention and treatment.

## 1. Introduction

Cannabis is one of the most widely used psychoactive substances in the United States, with approximately 43.5 million people over the age of 12 classified as users in the past year (Hasin and Walsh, 2020). Young adults exhibit the highest rates of cannabis use due to overlapping impulsivity and reward dysregulation that drives experimentation and susceptibility to addiction (Molina and Pelham, 2014). Cannabis use among young adults has risen in recently and is associated with negative consequences such as increased risk of injury and exacerbation of mental illness highlighting the need to address this growing public health problem (Crocker et al., 2021, Hamming and Jones, 2024; Mattingly et al., 2024).

Cannabis misuse is highly comorbid with ADHD, a neurodevelopmental disorder characterized by persistent hyperactive-impulsive and/or inattentional behaviors (American Psychiatric Association, 2016). For instance, 34-46% of adults (Notzon et al., 2020) and roughly one-third of adolescents (Zaman et al., 2015) in treatment for Cannabis Use Disorder (CUD) present with an ADHD diagnosis. These disorders share genetic overlap, with risk for cannabis use and CUD causally related to ADHD (Artigas et al., 2020; Nielsen et al., 2024). Substance use risk in ADHD has been linked to deficits in executive functioning that may drive use through impaired decision-making and inhibitory control. Another pathway proposed to link ADHD and cannabis use involves the effects of cannabis on neural systems underpinning ADHD symptomology (Gujska et al., 2023).

Cannabis use has been reported to have therapeutic effects on symptoms of inattention and hyperactivity/impulsivity (Dallabrida et al., 2024). Individuals with ADHD-related symptoms report using cannabis to alleviate symptoms (Mitchell et al., 2016; Hupli, 2018). These therapeutic effects may increase motivation to use cannabis among individuals with elevated ADHD symptoms, consistent with the self-medication hypothesis (Swanson et al., 2017). However, the neurobiological mechanisms linking ADHD symptomology, cannabis use, and neural function remain misunderstood.

Delta-9-tetrahydrocannabinol (THC) is the primary psychoactive compound found in cannabis, producing the feeling of being high through the endocannabinoid system by binding to CB1R, the G-protein coupled receptor (GPCR) most abundant in the human brain and locally encoded by the *CNR1* gene (Lu et al., 2008; Chayasirisobhon, 2020). The therapeutic effects reported in ADHD may be explained neurobiologically via the endocannabinoid system, which modulates small molecule neurotransmission throughout the brain, including the basal ganglia, cerebellum, striatum, nucleus accumbens, prefrontal cortex, and hippocampus, areas in which CB1R is densely expressed. THC has agonistic effects at CB1Rs, suppressing GABA release to increase local neurotransmitter efflux, including dopamine and norepinephrine (Laaris et al., 2011; Bloomfield et al., 2017). These are the primary neurotransmitters implicated in ADHD as commonly prescribed ADHD treatments block reuptake to increase synaptic levels in regions overlapping with CB1Rs (Aguiar et al., 2010; Faraone and Buitelaar, 2010; Mehta et al., 2019; Cho et al, 2024). While CB1R function is well understood at a cellular level, there is limited understanding regarding how CB1R interacts with neural substrates of ADHD and shapes large-scale network function (Ryan et al., 2024).

As legalization and access to cannabis increase, understanding neurobiological vulnerabilities for cannabis use and self-medication behaviors is crucial. We sought neural evidence of the hypothesis that cannabis promotes self-medication behaviors in relation to ADHD symptoms. We hypothesized that CB1 receptor localization would overlap with ADHD symptom-related functional deficits, offering a neural mechanism by which cannabis use could improve ADHD symptoms. We integrated *CNR1* gene expression from the Allen Human Brain Atlas with functional neuroimaging during a stop-signal task (SST), a probe of ADHD-related attentional and cognitive control deficits (Morita et al., 2016; Senowski et al., 2023). Here we show that SST hypoactivation in *CNR1*-expressing regions offers a mechanistic link between ADHD symptom severity and longitudinal cannabis use patterns in a non-clinical sample.

## 2. Materials and Methods

### 2.1 Participants

Data were collected from 466 participants aged 18-19 years old and in their first year of a four-year undergraduate program. Two samples of participants completed identical procedures except 165 (male=58, female=107) students completed an fMRI whereas 301 participants (male=90, female=211) only completed online procedures due to COVID-19 restrictions. The neuroimaging sample was recruited between 2018-2021 from undergraduate universities in the Chapel Hill, North Carolina area. The online sample was recruited nationwide through online platforms in the fall of 2020 with no exclusionary criteria beyond age and student status. Exclusion criteria for the neuroimaging sample were routine use of psychoactive medications, neurological disorders, psychiatric disorders other than past mood and anxiety disorders, baseline substance use disorders, and MRI contraindications. Presence of psychiatric disorders were screened for with a Mini-International Neuropsychiatric Interview (M.I.N.I.). A urine drug screen (Biotechnostix, Inc.) and alcohol breathalyzer test (FC-10, Lifeloc Inc.) were administered before the scan. Cocaine, cannabis, opioids, amphetamines, and methamphetamines were screened, with no participants testing positive. ADHD was not a criterion for inclusion or exclusion for either sample. Further information regarding study participants is presented in Table 1.

**Table 1.** Sample characteristics and descriptive statistics for self-report data. Table 1: Self-reported characteristics for entire sample (n=465) and each cannabis use class. CAARS: Conners Adult ADHD Rating Scales. AUDIT: Alcohol Use Disorder Identification Test. Cannabis use defined by the number of times used in the year prior to each timepoint. Percentages are defined by lifetime use prior to the timepoint.

| Variable | Total Sample |  |  | Non-User Class |  |  | Low User Class |  |  | High User Class |  |  |
| --- | --- | --- | --- | --- | --- | --- | --- | --- | --- | --- | --- | --- |
|  | N (%) | Mean | Std. Dev. | N (%) | Mean | Std. Dev. | N (%) | Mean | Std. Dev. | N (%) | Mean | Std. Dev. |
| Male | 31.80% |  |  | 38.80% |  |  | 23.30% |  |  | 28.70% |  |  |
| Female | 68.20% |  |  | 61.20% |  |  | 76.70% |  |  | 71.30% |  |  |
| CAARS DSM-IV: ADHD Symptoms Total |  | 15.4 | 9.8 |  | 14.1 | 9.3 |  | 15 | 9.8 |  | 18.8 | 10.2 |
| CAARS DSM-IV: Inattentive Symptoms |  | 7.8 | 5.5 |  | 7 | 5.2 |  | 7.7 | 5.6 |  | 9.6 | 5.9 |
| CAARS DSM-IV: Hyperactive-Impulsive Symptoms |  | 7.7 | 5.2 |  | 7.1 | 5.1 |  | 7.4 | 4.9 |  | 9.3 | 5.4 |
| Cannabis Use at Baseline | 42.50% | 14.4 | 77.5 | 0% | 0 | 0 | 53.40% | 2.4 | 6.9 | 90.10% | 62.2 | 156.9 |
| Cannabis Use at First Follow-up | 50.00% | 13.8 | 47.5 | 0% | 0 | 0 | 69.90% | 2.7 | 5.9 | 98.00% | 69.5 | 92.6 |
| Cannabis Use at Second Follow-up | 56.70% | 25.4 | 96.4 | 0% | 0 | 0 | 88.40% | 3.8 | 6.2 | 99.00% | 149.5 | 202.7 |
| Cannabis Use at Third Follow-up | 59.60% | 20.8 | 59.3 | 0% | 0 | 0 | 96.60% | 6 | 9.8 | 100.00% | 123.5 | 107.2 |
| AUDIT Score at Baseline |  | 3.4 | 4.1 |  | 1.6 | 2.5 |  | 3.8 | 3.8 |  | 6.6 | 5 |
| AUDIT Score at First Follow-up |  | 4.0 | 4.2 |  | 1.7 | 2.7 |  | 4.7 | 3.9 |  | 7.3 | 4.7 |
| AUDIT Score at Second Follow-up |  | 4.6 | 4.1 |  | 2.6 | 3 |  | 5.1 | 3.6 |  | 7.4 | 5.1 |
| AUDIT Score at Third Follow-up |  | 4.6 | 3.7 |  | 2.2 | 2.2 |  | 4.8 | 3.3 |  | 8.2 | 6.1 |

Growth mixture models (GMMs) included 466 participants to classify cannabis use patterns. Sixteen participants missing baseline questionnaire data were excluded from mediation models, resulting in an analytic sample of 450 participants. Due to technical difficulties (e.g. scanner reboot or task malfunctioning), fMRI data was not available for 21 participants, leaving 144 participants contributing data to fMRI analyses. Questionnaire data from these 21 participants were included in all other analyses.

### 2.2 Procedures

Upon screening eligible for study participation, participants gave electronic informed consent in accordance with University of North Carolina Chapel Hill Office of Human Research Ethics guidelines. Subjects remotely completed self-report questionnaires in REDCap (Harris et al., 2009) at baseline and three yearly remote follow-up surveys. Participants in the neuroimaging sample underwent an fMRI scan including a SST (Figure 1A).

**Figure 1.**
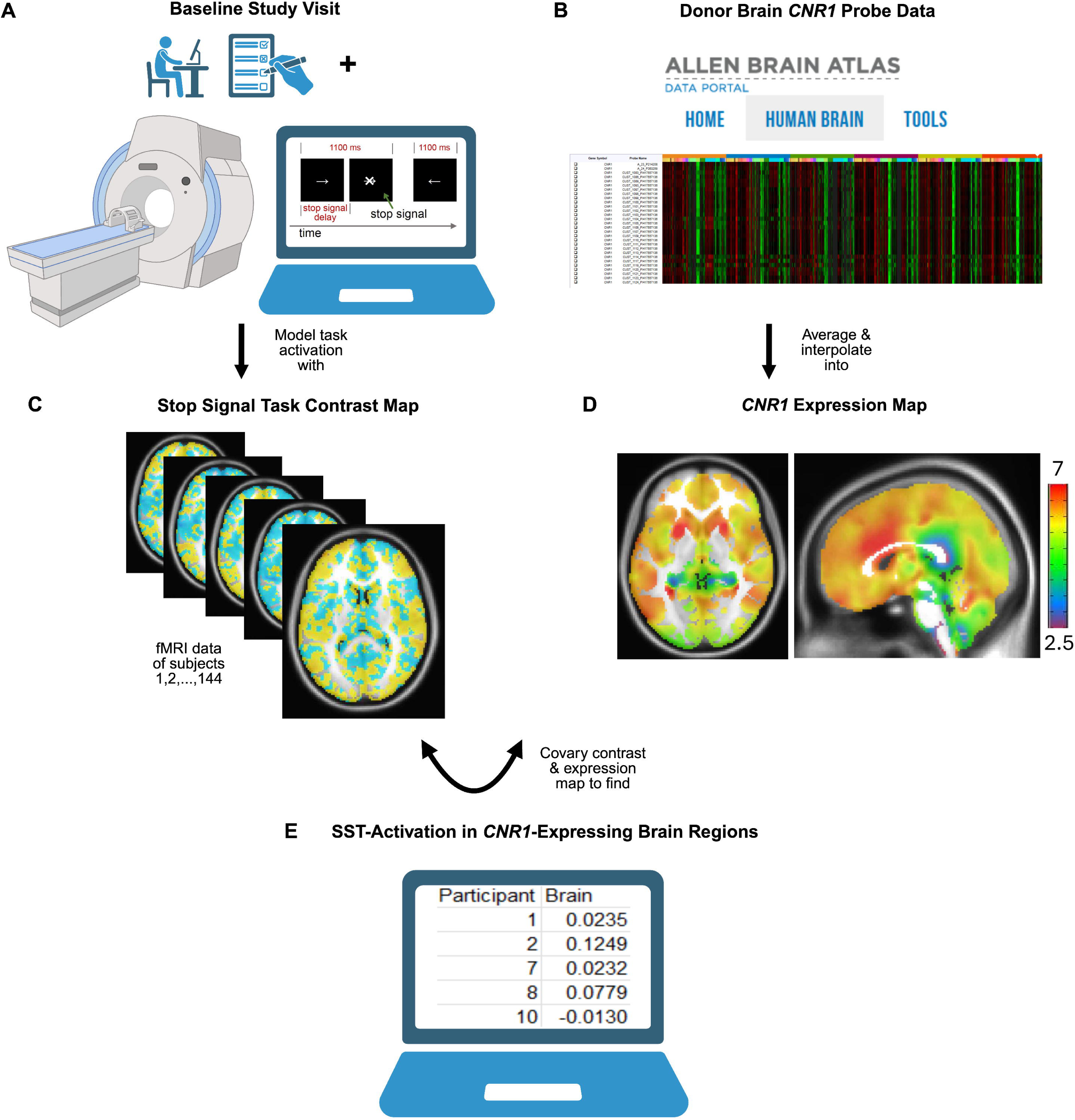
Mapping functional activation in neural areas of CB1R Activation. A) Baseline Study Visit: Participants completed questionnaires (ex. CAARS, CDDR, etc.) and underwent an fMRI scan in which they completed a stop signal task. B) Microarray data from two human full-brain donors were downloaded from the AHBA. Probes measuring *CNR1*-expression were extracted, averaged, and interpolated into a 3D MNI space to map *CNR1*-expression into 3D space. C) Contrast maps of successful stops vs go trials were created for each participant’s stop signal task data to represent activation during inhibitory control. D) *CNR1*-Expression Map. Axial cut at position z=0, sagittal cut at x=2. Activation quantified by the scale provided, red representing high gene expression (≥7 log_2_-intensity) ranging to purple for reduced expression (≤2.5)). E) For each participant, the covariance of this map with their SST contrast maps was calculated. The covariance measure indicates brain function in regions with higher-versus-lower *CNR1*-expression.

**Figure 2.**
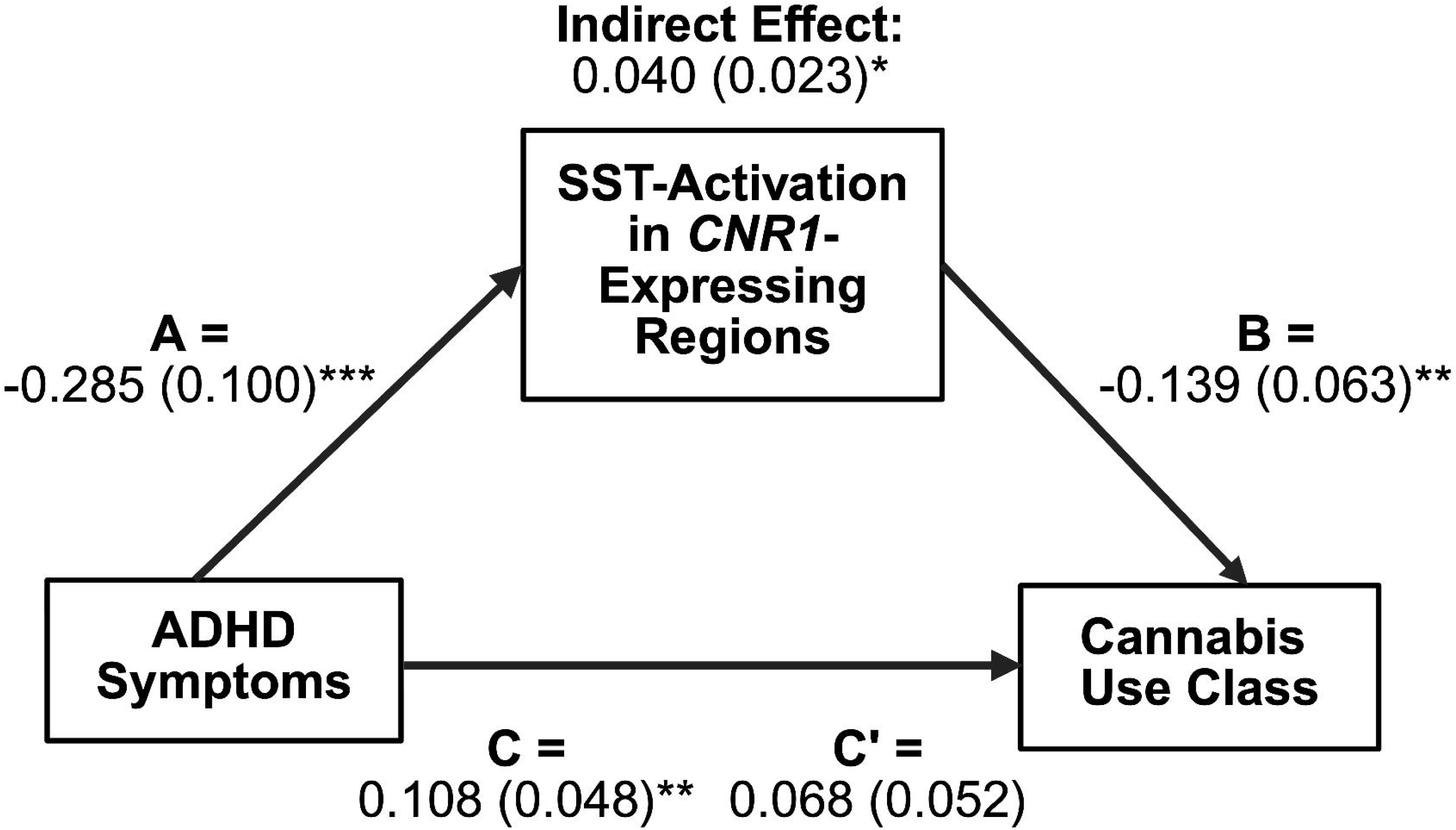
*CNR1* Gene Expression in the Stop Signal Task. A) *CNR1*-Expression Map. Gene expression microarray probe data from the AHBA was extracted and interpolated into a 3D volume in MNI space. Axial sections from inferior to superior, activation quantified by the scale provided, red representing high gene expression (≤7) ranging to purple for reduced expression (≥2.5). B) Functional activation during the SST in *CNR1*-expressing regions by cannabis use class. Covariance between activation and cannabis use class revealed a trend toward significance (*p*=0.085) High users showed deficits in regions overlapping with *CNR1* receptor expression. \**p*<0.10, \*\**p*<0.05. C) Scatter plot indicating that greater ADHD symptoms are correlated with reduced activation in brain regions overlapping with higher *CNR1*-expression (*r*=-0.23, *p*=0.007).

This study integrates self-reported ADHD symptoms, fMRI data from a SST, *CNR1* transcriptomic data from the Allen Human Brain Atlas, and longitudinal cannabis use trajectories. Finally, a mediation analysis tested whether the effects of ADHD symptoms on cannabis use are mediated through SST-related brain function in *CNR1*-expressing regions.

### 2.3 Self-Report Data

Substance use patterns were assessed via the Customary Drinking and Drug Use Record (CDDR) at baseline and each follow-up (Brown et al., 1998). Participants provided information regarding alcohol and cannabis use. Cannabis use frequency in the past year was assessed at each time point with the question, “In the last year, how many times have you used marijuana?”. Current cannabis use was quantified with, “When was the last time you used marijuana (# of days ago)?”.

The Conners Adult ADHD Rating Scales (CAARS) questionnaire was administered at baseline to measure presence and severity of ADHD symptoms (Connors et al., 1999; Kooj et al., 2013). Questions reflected DSM-IV diagnostic criteria for ADHD with participants indicating symptom frequency with a 4-point Likert-type scale regarding inattentive, hyperactive-impulsive, and total symptoms for each participant. The Total Symptoms subscale was used in analyses due to likely associations of attentional and impulsive symptoms with cannabis use (Hernandez and Levin, 2024).

The Adverse Childhood Experiences (ACE) questionnaire surveyed exposure to childhood adversity before age 18 (Felitti et al., 1998; Zarse et al., 2019). Questions assessed household substance misuse and other childhood experiences that may affect adult well-being. Given the associations of a score of four or more with substance misuse (Hines et al., 2023) this cutoff was used as a binary covariate.

The MacArthur Scale of Subjective Social Status (Adler et al., 2000; Galvan et al., 2023) measures socioeconomic status. Family income during adolescence was ranked through a categorical income scale, using a range of binned incomes and the options “Don’t know” and “No response”.

### 2.4 Stop-Signal Task

The fMRI SST measures brain activity during response inhibition, or the ability to suppress a prepotent motor response. SSTs have been used to assess ADHD-related deficits in inhibitory control related to inattentive and hyperactive-impulsive symptoms. The SST engages multiple neural networks supporting attentional processes including stimulus detection, context monitoring, sustained vigilance, and flexible allocation of attentional resources (Chikazoe et al., 2009; Aron et al., 2014). The task included 300 trials, with go trials requiring a rapid button press for directional arrows and 25% (75) of trials including a stop signal indicating participants should withhold their response. The stop signal occurred after an adjusting stop-signal delay to enable successful stops in approximately 50% of stop trials. Further information regarding task parameters and behavioral performance can be found in *Supplementary Materials* (Table S1) and our prior publication (Elton et al., 2023).

### 2.5 fMRI Data Acquisition, Preprocessing, and Task Activation Analysis

Data acquisition, image preprocessing, and modeling for this study have been previously reported [40], and further information can be found in *Supplementary Materials*. We used the contrast of successful stop versus correct go trials to isolate inhibitory control.

### 2.6 Cannabinoid Receptor Mapping

The Allen Human Brain Atlas contains publicly available quantitative maps of transcript distribution across donor brains to expand knowledge regarding human transcriptional architecture. Approaches integrating donor tissue with functional neuroimaging have been previously used to characterize gene-behavior relationships (Salvan et al., 2022). The dataset contains 2 bilateral, clinically unremarkable, male donor brains from individuals aged 24 and 39 years old. Donors were selected for analysis due to highly correlated expression across anatomical structures and bilateral data availability (Hawrylycz et al., 2012).

The *CNR1* gene locally encodes the CB1R protein, the GPCR which modulates neural synaptic transmission in response to cannabinoids (Lu and Mackie, 2021). Additionally, *CNR1* was selected due to its prior genotypic association with cannabis dependence, trait impulsivity, and ADHD susceptibility (Lu et al., 2008; Agarwal et al., 2009; Bidwell et al., 2013).

Quantitative normalized microarray probe data for both donors were extracted (Figure 1B) and averaged to estimate local gene expression, allowing for evaluation of associations across the full range of probe expression (French and Paus, 2015; Wagstyl et al., 2024). Probe expression values represent log_2_-transformed, normalized microarray hybridization intensities (dimensionless). Three-dimensional standardized anatomical MNI coordinates are provided for each site of gene expression to allow for mapping of transcriptomic data alongside imaging data. Using MATLAB R2023b (Niazai et al., 2023), we averaged expression values across donors and interpolated them into a whole brain 3D volume in MNI space using the natural neighbor approach (Wagstyl et al., 2024). This map was resampled to match the 2mm voxel size of the contrast maps and masked with the group-derived binary gray matter mask.

### 2.7 Gene Expression-Neuroimaging Covariance Analysis

To examine the relationship between patterns of activation during the fMRI SST and the endocannabinoid system, we integrated the interpolated *CNR1* map with participants’ whole-brain SST contrast maps (Wagstyl et al., 2024). Specifically, we created a 1D vector of voxel values for both the contrast (Figure 1C) and expression maps (Figure 1D). To minimize outlier effects, SST vector values were Winsorized using the 1^st^ and 99^th^ percentile values. For each participant, the covariance between *CNR1* values and SST contrast values was calculated to index neural activation in regions with higher-versus-lower *CNR1*-expression (Arnatkeviciute et al., 2019; Seidlitz et al., 2020) (Figure 1E). The covariance value for each participant was utilized as quantitative brain-related mediator variable.

### 2.8 Growth Mixture Model

Prior research analyzing cannabis use patterns over time has shown that users can be separated into groups based on frequency of use, sharing similar clinical characteristics, risk factors, and psychosocial outcomes (Hurd et al., 2014; Kosty et al., 2016). Growth mixture modelling is often used for statistical analysis of longitudinal data to reveal latent classes (“class”) with distinct developmental trajectories (Greenwood et al., 2019).

Cannabis use was highly variable across individuals in our sample across time (Figure S1). Mplus Version 8.11 (Muthen and Muthen, 2017) was used to create a GMM (Figure S2) including only cannabis users to determine latent classes of cannabis use from yearly reports of use frequency within the past year. To limit outlier effects, frequency was truncated to 365 times per year, representing the 99^th^ percentile at each timepoint. Cannabis use were modelled using a negative binomial distribution to account for overdispersion and the effect of contagious events. The separate study samples were accounted for with a binary covariate. Non-cannabis users (n=219) were defined as reporting no lifetime use at baseline and no cannabis use at any follow-up period; these participants were manually placed into a separate class.

As a part of the class enumeration process, several models with differing numbers of latent classes were computed (n=5) and contrasted by comparative model fit (−2 Log Likelihood, BIC, ABIC) and predicted class proportions to determine the most representative number of classes (Roberts et al., 2023) (Table S2). Class membership was utilized as an outcome variable in the mediation model (Hammerton et al., 2024).

### 2.9 Mediation Model

Mplus 8.11 was used to estimate a mediation model to examine whether function in *CNR1*-expressing brain regions linked ADHD symptoms to cannabis use patterns. The predictor (*x*) variable was the Total ADHD Symptoms subscale score of the CAARS, the categorical outcome variable (*y*) was the cannabis use class membership derived from the GMM, and the mediator variable (*m*) was SST activation in *CNR1*-expressing brain regions.

Self-reported sex assigned at birth was included as a covariate through binary coding of males as 0 and females as 1. To control for possible effects of recent cannabis use on SST activation, a binary variable indicated whether participants reported using cannabis in the past week at baseline (≤7 days prior=1, ≥8=0) (Leung et al., 2020). To account for the high co-use of cannabis and alcohol, a binary covariate indicated if participants reported lifetime alcohol use at baseline (Yes=1, No=0) (Leung et al., 2017; Yurasek et al., 2017). The effect of socioeconomic status on cannabis use (Martin, 2019) was represented by a binary median-split variable of self-reported household income during adolescence (≥$100,000 annually=1, ≤$99,999=0). Participants that selected “Don’t Know” or “No Response” to this question (n=96), were accounted for with a missingness indicator variable in which responses were coded as 1, and other participant responses were coded as 0. The effect of adverse childhood experiences was accounted for through a binary covariate indicating adverse life experiences prior to the age of 18 (≥4=1, <4=0).

We also examined the possibility that the mediating relationship may be better explained by the reverse direction, i.e., that cannabis use alters the brain, resulting in ADHD symptoms. A secondary mediation model was run using baseline cannabis use as the predictor and ADHD symptoms as the outcome, keeping the same mediator variable (Figure S3).

## 3. Results

### 3.1 Self-Report Data

Self-reported data is recorded in Table 1.

### 3.2 Stop-Signal Task

Group-level SST activation and behavioral results have already been published (Elton et al., 2023). Significant positive activations were predominately found in bilateral prefrontal regions, anterior cingulate cortex, and visual cortex, whereas decreased activity was in primary and supplementary motor regions and ventromedial prefrontal cortex.

A supplemental analysis testing relationships between CAARS ADHD symptom subscales and whole brain voxel-wise SST activation identified no clusters that survived multiple comparison correction. There was a negative correlation between participant CAARS Total ADHD symptoms and the average activation in significant task-positive brain regions (*r*=-0.30, *p*<0.001; Figure S2), suggesting ADHD symptoms were associated with deficits in engagement of inhibitory control-related brain regions.

An additional supplemental analysis was run to determine the effects of head motion on fMRI results. Framewise displacement was calculated for each cannabis class and a one-way ANOVA showed head motion did not significantly predict cannabis use class (*F*(2,141)=0.43, *p*=0.649).

### 3.3 Gene Expression-Neuroimaging Covariance Analysis

The average covariance between participant’s fMRI SST contrast map and the *CNR1*-expression map (Figure 3A) was significantly greater than 0 based on a 1-sample t-test (*t*(143)=5.17, *p*<0.001, 95% CI[0.0134, 0.0299]), indicating significant SST activation in regions with higher *CNR1*-expression. The covariance was then examined by cannabis user class with an ANCOVA controlling for the covariates used in the mediation model (*F*(2,133)=2.52, *p*=0.085; Figure 3B). Post hoc comparisons indicated that compared with non-cannabis users, high users showed significantly reduced SST activation in *CNR1*-expressing regions (β=0.040, SE=0.018, *t*(132)=2.24, *p*=0.027). Low users showed a trend towards higher activation compared to high users (β=0.033, SE=0.017, *t*(132)=1.88, *p*=0.062).

**Figure 3.**
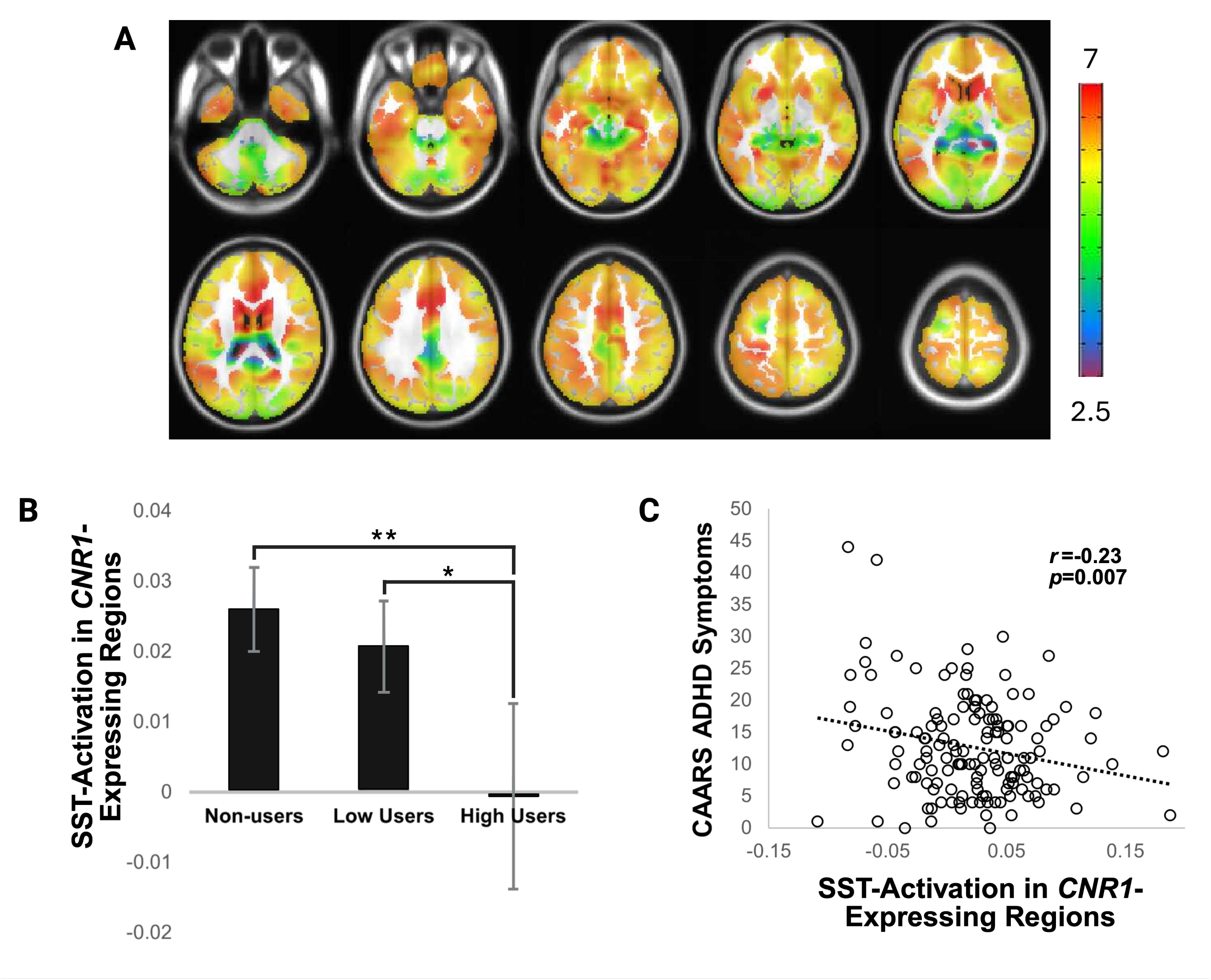
Model of ADHD symptoms predicting cannabis use class, including mediating effects of neural activation in *CNR1*-expressing brain regions. Relationships represented by a, b, and c paths were significant, and there was a trend (*p<*0.10) for an indirect effect in which ADHD symptoms related to cannabis use through deficits in brain function. \**p*<0.10, \*\**p*<0.05, \*\*\**p*<0.005.

Greater ADHD symptoms correlated with reduced covariance between SST activation and *CNR1*-expression (*r*=-0.23; *p*=0.007; Figure 3C), indicating reduced activation in regions with higher *CNR1*-expression.

### 3.4 Growth Mixture Model

Model fit indices and estimated class proportions for each GMM latent class model (n=5) are provided in Table S2. Our results indicated that there were 2 distinct latent classes of cannabis use within our subsample of users. More specifically, the 2-class model presented the lowest BIC and ABIC comparative fit indices (BIC=4099.858; ABIC=4058.648). While the loglikelihood value was not the lowest (LL=-2014.118) it presented an elbow in the data, indicating an acceptable model fit. The 2-class model also had the most proportionate estimated distributions of participants across classes (Table S2). Based on these results, we labelled the classes as “non-users” (n=219), “low users” (n=146), and “high users” (n=101). This classification of cannabis use trajectories aligns with previous literature categorizing substance use and related outcomes (Grevenstein and Kroninger-Jungaberle, 2015). Further descriptives of each class are detailed in Table S2.

### 3.5 Mediation Model

The mediation model examining the relationship between ADHD symptoms, cannabis use patterns, and functional neural activation (Figure 4), demonstrated a good model fit: X^2^(17)=0.000, *p*=0.000; CFI=1.000; TLI=0.000; RMSEA=0.000; SRMR=0.000.

In the *a* path, greater ADHD symptom severity significantly predicted less covariance between SST-activation and *CNR1*-expressing regions (β=-0.285, SE=0.100, *p*=0.004). In the *b* path, lesser covariance between SST-activation and *CNR1*-expression significantly predicted a greater cannabis use class membership (β=-0.139, SE=0.063, *p*=0.028; Table 2).

**Table 2.**
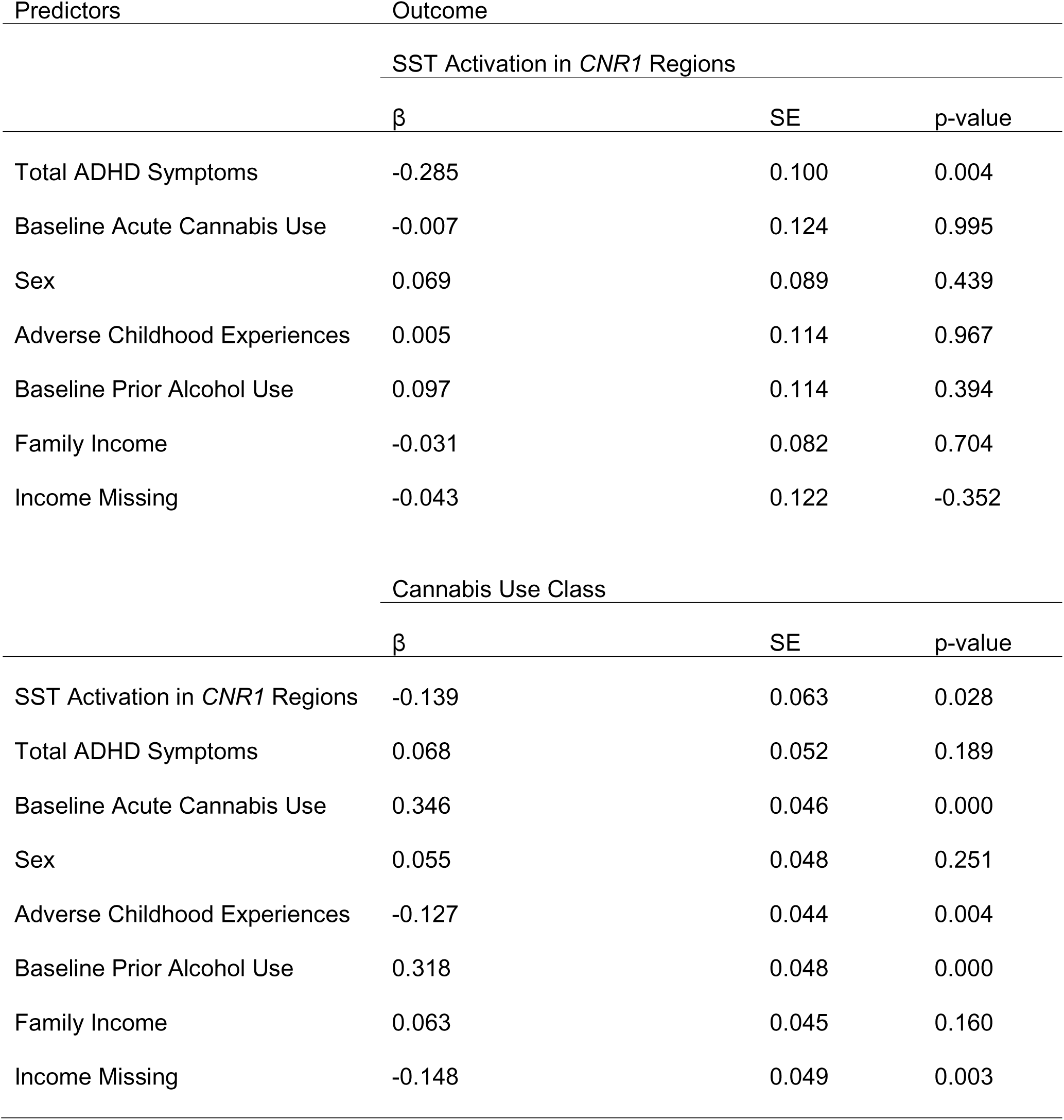
Standardized Mediation Model Resul. Table 2: Standardized mediation model results including all variables and covariates. Total ADHD symptoms are defined by the DSM-IV Total ADHD Symptoms subscale of the CAARS questionnaire. SST-activation in *CNR1* regions is quantified from the covariance of participant SST contrasts and *CNR1* expression maps. Baseline acute cannabis use is defined as the self-reported number of times used within 7 days prior to the baseline study visit binarized as number of times used <8=0 and >7=1. Sex is binarized as female=1 and male=0. Adverse childhood experiences are defined as having ≥4=1 and <4=0. Family income is binarized based on a median split of $100,000+=1 and below=0, and income missing is a measure of “Don’t know” and “I don’t know” responses to the MacArthur SES Ladder of parental income to isolate family income effects. Cannabis use class is coded as 1=non-user group, 2=low user group, 3=high user group.

| Predictors | Outcome |  |  |
| --- | --- | --- | --- |
|  | SST Activation in *CNR1* Regions |  |  |
| | $\beta$ | SE | p-value |
| Total ADHD Symptoms | -0.285 | 0.100 | 0.004 |
| Baseline Acute Cannabis Use | -0.007 | 0.124 | 0.995 |
| Sex | 0.069 | 0.089 | 0.439 |
| Adverse Childhood Experiences | 0.005 | 0.114 | 0.967 |
| Baseline Prior Alcohol Use | 0.097 | 0.114 | 0.394 |
| Family Income | -0.031 | 0.082 | 0.704 |
| Income Missing | -0.043 | 0.122 | -0.352 |
|  | Cannabis Use Class |  |  |
| | $\beta$ | SE | p-value |
| SST Activation in *CNR1* Regions | -0.139 | 0.063 | 0.028 |
| Total ADHD Symptoms | 0.068 | 0.052 | 0.189 |
| Baseline Acute Cannabis Use | 0.346 | 0.046 | 0.000 |
| Sex | 0.055 | 0.048 | 0.251 |
| Adverse Childhood Experiences | -0.127 | 0.044 | 0.004 |
| Baseline Prior Alcohol Use | 0.318 | 0.048 | 0.000 |
| Family Income | 0.063 | 0.045 | 0.160 |
| Income Missing | -0.148 | 0.049 | 0.003 |

The total effect of ADHD symptoms on cannabis use class (c path of Figure 4) was significant (β=0.108, SE=0.048, *p*=0.026) while the direct effect was not (c prime path of Figure 4; β=0.068, SE=0.052, *p*=0.189), indicating that the total effect is partly driven by the mediator variable. The indirect effect of total ADHD symptoms on cannabis use class indicated a trend and partial mediation towards ADHD symptoms predicting a higher use class membership through the mediator variable (β=0.040, SE=0.023, *p*=0.083).

In the reverse model (Figure S3) there was no significant relationship between baseline cannabis use and covariance between SST-activation with *CNR1-*expression (*a* path). As with the *a* path in the original model, relationship between the brain covariance measure and ADHD symptoms was significant, representing the *b* path in the reverse model (Table S3).

## 4. Discussion

This four-year longitudinal study incorporating fMRI, gene expression data, and longitudinal measures identified evidence linking ADHD symptoms to increased cannabis use through in *CNR1*-expressing brain regions. We examined whether brain activation in *CNR1*-expressing regions during a SST mediated the association between ADHD symptoms and cannabis use trajectories. The mediation analysis provided preliminary support for our hypothesis that ADHD symptom severity was associated with reduced activation during the SST in *CNR1*-expressing regions, which in turn predicted a greater likelihood of belonging to a higher-use class. These results support Positron Emission Tomography (PET) imaging associations between lower in-vivo CB1R availability and impulsive novelty-seeking traits that play a key role in ADHD pathogenesis (Ceccarini et al., 2015; Miederer et al., 2026). The analysis focused on dimensional ADHD symptoms rather than categorical ADHD diagnosis, suggesting these relationships are relevant across the continuum of symptoms. These findings implicate *CNR1*-enriched circuits involved in attention and impulsivity as a pathway through which ADHD symptomology contributes to long-term patterns of cannabis use.

Reduced SST-related activation in individuals with greater ADHD symptom severity aligns with established inhibitory and attentional control deficits in ADHD (Rubia, 2018). Decreased recruitment of right inferior frontal, pre-supplementary motor, and striatal regions consistent with prior ADHD neural phenotypes (Aron et al., 2014). Our analysis extends this literature by demonstrating these deficits in regions with higher *CNR1*-expression, suggesting altered endocannabinoid signaling may contribute to these functional differences. Since *CNR1* encodes CB1Rs, widely expressed in areas of inhibitory control and modulators of neurotransmitter release, reduced SST-activation may reflect disrupted endocannabinoid modulation of inhibitory synapses in ADHD. This observation coincides with prior genetic evidence linking *CNR1* to ADHD susceptibility and impaired inhibition. By leveraging transcriptomic maps of *CNR1* expression, we localized these functional deficits to molecularly defined regions, bridging gene-level and system-level processes.

Prior neuroimaging studies have reported greater alterations in inhibitory control networks and reduced CB1R availability in the cortex and striatum following chronic cannabis exposure (Lorenzetti et al., 2020; Hirvonen et al., 2012). This dysregulation may impair top-down cortical control and increase impulsive decision-making, increasing susceptibility to further cannabis use. The observed stepwise decline in SST-activation across non-, low-, and high-use groups mirrors *CNR1*-expression covariance, supporting a dose-dependent relationship between inhibitory network engagement and cannabis consumption. This gradient could reflect cannabis-induced neuroadaptations in which individuals with more prior use exhibit baseline functional deficits in CB1R-rich inhibitory regions (Tseng and Molla, 2025), potentially leading to elevated ADHD symptoms. A mediation analysis testing this reverse model did not support a pathway in which baseline cannabis use predicted neural activation, in turn predicting ADHD symptoms (Figure S3; Table S3). We found no significant paths except between SST activation and ADHD symptoms suggesting a robust linkage of neural activation with ADHD severity and no causal pathway from early cannabis use to altered network function. Although recreational and chronic cannabis use are associated with cognitive deficits and bidirectional neurodevelopmental interactions cannot be ruled out, the current evidence favors pre-existing neural differences mediating ADHD-cannabis associations rather than prior cannabis use (Kelly et al., 2017; Rasmussen et al., 2017; Burggren et al., 2020).

These findings have translational relevance for the self-medication hypothesis. Individuals with elevated ADHD often report using cannabis to affect dysregulated arousal, yet cannabis use may exacerbate neural deficits in inhibitory control via CB1R downregulation or desensitization, perpetuating a maladaptive feedback loop. Chronic hypoactivation of endocannabinoid system-linked inhibitory circuitry may compromise behavioral regulation, predisposing individuals to increased cannabis consumption. Hypoactivation within this network could reflect reduced cognitive control and altered reward sensitivity, explaining why ADHD symptomology predicts impaired behavioral regulation and higher substance use in young adulthood (Kim-Spoon et al., 2016). Currently, there are no approved pharmacotherapies for CUD, particularly in the context of comorbid substance use and ADHD (Connor et al., 2021). Linking gene-expression maps with neural activation and behavioral phenotypes provides a framework for mechanistic neuroscience.

### 4.1 Limitations

Several limitations should be considered when interpreting the results. ADHD symptoms were assessed using a single baseline CAARS self-reported measure, which may not capture fluctuations across the 4-year period. Despite prior validation (Christiansen et al., 2012), the CAARS self-report format has been criticized for overestimating symptom severity relative to structured clinical diagnoses. Future work should incorporate repeated, multi-informant assessments to more precisely characterize ADHD symptoms. Although the SST is used to probe ADHD-related deficits, it primarily indexes response inhibition. Additional paradigms such as the Continuous Performance Task may be better suited to isolate other attentional processes implicated in ADHD. Future work can examine hyperactive-impulsive and inattention subtypes, allowing for more precise mapping of mechanisms to specific symptom domains, informing individual-level treatment approaches. The *CNR1*-expression map was extracted from normative post-mortem data and does not capture interindividual variability in CB1 receptor density; future integration of PET imaging would provide direct receptor validation. Additionally, multiband fMRI sequences like the one used in the current study have reduced signal in subcortical regions, which could have affected our power to detect associations with *CNR1*-expression. Latent class models capture use patterns but do not reflect potency, route of administration, polysubstance use, or early-life initiation, all of which may moderate neural outcomes (De Jonge et al., 2022). The restricted fMRI subsample (n=144) and modest analytical sample size (n=450) reduce power and constrain sensitivity to detect small effects [71]. Although the model controlled for sex, preclinical data shows sex-specific variation in *CNR1*-expression (Castelli et al., 2014), warranting future exploration.

### 4.2 Conclusions

These findings provide evidence for a CB1 receptor-related pathway linking ADHD symptoms to cannabis use trajectories. Individuals with greater ADHD symptom severity showed reduced engagement of *CNR1*-rich regions during response inhibition, and this lower activation predicted heavier cannabis use across four years. Although the indirect effect was modest, the pattern is consistent with a preliminary mechanistic mediation model. Ultimately, receptor-informed models can guide early identification of at-risk individuals and the development of targeted interventions for cannabis use disorder in populations with neurodevelopmental vulnerabilities.

## Funding

This work was supported by NIH grant K01AA026334 to AE. Funding sources did not have involvement in study design, collection, analysis, or writing of the report for publication.

## Conflict of Interest

The authors declare no conflicts of interest.

## Author Contributions

JA – Conceptualization, Methodology, Formal Analysis, Writing – original draft; SC – Supervision, Writing – review and editing; JHA – Investigation, Writing – review & editing; CCY – Visualization; SJN – Writing – review and editing; AE – Supervision, Investigation, Writing – review & editing.

## Data Availability

All data produced are available online at https://github.com/amanda-elton-lab/Neural-Correlates-of-ADHD-as-Phenotypes-of-Cannabis-Use.git

## Acknowledgements

We thank the participants for their contribution to this study and acknowledge the Allen Institute for Brain Science for making the Allen Human Brain Atlas data publicly available. The authors acknowledge the role of Dr. Charlotte A. Boettiger in the earlier phases of this project and for her mentorship to AE on K01AA026334.

## Ethical Approvals

This study was approved by the Institutional Review Board at the University of North Carolina Chapel Hill (neuroimaging sample: IRB18-081 and online sample: IRB20-1398). Written informed consent was obtained from all participants prior to their inclusion in the study.

## Data Availability Statement

Code used for sections 2.7 and 2.9 are publicly available on GitHub (https://github.com/amanda-elton-lab/Neural-Correlates-of-ADHD-as-Phenotypes-of-Cannabis-Use.git).

## Notes

### Competing Interest Statement

The authors have declared no competing interest.

