## Supplemental for "ADHD Symptoms and Cannabis Use: The Role of Cannabinoid Receptor 1 and Neural Response Inhibition"

**Supplementary Materials**

1. *Stop-Signal Task*

In this paradigm, the SST was presented to participants in the neuroimaging sample using PsychoPy software (Peirce, 2007; Figure 1A). The stop signal task used was divided into two consecutive runs of approximately 5.5 minutes each for a total of 300 trials. Each run included three 100 second blocks of 50 trials neighbored by four 12 second blocks of a fixation cross. In each trial, an arrow appeared in the center of the screen facing either left or right. Participants were given a button box in the scanner and instructed to make a left or right button press corresponding to the arrow direction presented on the screen. In 25% of the trials (n=75), after a brief delay an “X”, also referred to as a stop-signal, was presented on top of the on-screen arrow. The delay before the “X”, referred to as the stop-signal delay, started at 200ms and was adaptively shortened by 50ms after a failure to withhold a response and lengthened by 50ms after each successful stop. The stop-signal delay could only reach a maximum of 900ms. Before starting the task, participants were instructed to withhold their response if the “X” were to appear. In 25 of the trials (8%), a bidirectional arrow that did not require a button press appeared to assist in deconvolving blood oxygenation level-dependent (BOLD) signals. Before entering the scanner, participants underwent a 50-trial practice and were reminded of the importance of speed, accuracy, and not anticipating the stop-signal during the task.

The following behavioral performance indices were computed: (1) go accuracy: the proportion of correct responses on go trials; (2) go reaction time (go RT): the mean RT for correct go trials; (3) stop accuracy: the proportion of successful inhibition on stop trials; (4) stop signal delay (SSD), defined as the average delay between the onset of the go stimulus and the onset of the stop signal and (5) stop signal reaction time (SSRT) was used to quantify the latency of successful motor inhibition based on the Horse-Race Model (Logan et al., 1984; Logan & Cowan, 1984). The integration method, with replacement of go omissions by the maximum RT, was used to calculate SSRT (Verbruggen et al., 2019). SSRT was calculated by subtracting the mean SSD from the nth percentile of the go RT distribution, where n represents the probability of responding on stop trials (i.e., stop error rate). RTs > three standard deviations from the individual mean within each block were excluded. The performance is summarized in the Supplementary Table 1.

2. *fMRI Data Acquisition, Preprocessing, and Task Activation Analysis*

During the SST, fMRI data was collected using multiband echo-planar imaging (EPI) and a 32-channel head coil (Siemens Healthineers, Erlangen, Germany) on a Siemens 3Tesla Prisma scanner. Sequence information for acquiring BOLD images were multiband factor=8, TR =800ms, TE=37ms, flip angle=52°, 2mm isotropic voxels, 72 sagittal slices, field of view (FOV)=208x208, bandwidth=2290 Hz/pixel, interleaved acquisition. An Anterior-to-Posterior (AP) phase encoding direction was used for the first 413 TRs, followed by a phase encoding for the last 413 TRs for a total scan time of about 11 minutes. An update to the scanner occurred for the final 27 subjects, therefore an adjusted sequence with the bandwidth=2186 Hz/pixel and TE=38.2ms was used for these participants.

To assist with tissue segmentation and alignment, a T1-weighted (T1w) magnetization-prepared rapid gradient echo (MPRAGE) was acquired with sequence parameters of a TE=2.3ms, TR=2530ms, 1mm isotropic voxels, flip angle=9°, FOV=256x265, for a total of 176 sagittal slices.

Brain images collected during the T1w and stop-signal task scans were processed using fMRIPrep (Esteban, 2019). Preprocessing for T1-weighted images included skull-stripping, tissue segmentation and reconstruction, volume-based spatial normalization, and estimation. BOLD fMRI data preprocessing included slice-timing correction, motion estimation and realignment, registration to the T1 image, confounds estimation, and normalization to Montreal Neurological Institute (MNI) standard space.

First level analyses were modeled and estimated using 3dDevconvolve and 3dREMLfit in AFNI software (version 20.3.00) utilizing the general linear modelling approach (Friston, 1994). Voxel-wise activation during the stop-signal task was modelled by contrasting activation during successful stops (withheld response when the stop-signal appeared) with correct go trials. Contrast maps of successful stops minus go trials were created for each participant with valid fMRI data (n=144), resulting in a single brain map indexing cognitive function during the task for each participant. A sample-derived binary grey matter mask was applied to each contrast map.

Further acquisition, preprocessing, and task analysis information for this study can be found in the previous report (Elton et al., 2024).

**Supplemental Tables**

Table S1. Stop Signal Task Performance Descriptive Statistics

| Variables | Mean (SD, *n* = 144) |
| --- | --- |
| Go accuracy (%) | 97.25 (5.97) |
| Go reaction time (RT, ms) | 747.14 (181.67) |
| Stop accuracy (%) | 40.83 (7.37) |
| Stop signal delay (SSD, ms) | 532.94 (182.23) |
| Stop signal reaction time (SSRT, ms) | 255.32 (64.04) |

Table S2. Growth Mixture Model Class Fit Statistics

| Model | Log Likelihood | BIC | ABIC | Class Proportions |
| --- | --- | --- | --- | --- |
| 1 Class | -2201.301 | 4463.205 | 4428.335 | 1.000 |
| 2 Class | -2014.118 | 4099.858 | 4058.648 | 0.632,0.377 |
| 3 Class | -2003.915 | 4101.489 | 4047.599 | 0.332,0.534,0.134 |
| 4 Class | -2003.429 | 4122.556 | 4055.986 | 0.130,0.308,0.008,0.555 |
| 5 Class | -2000.677 | 4139.090 | 4059.840 | 0.150,0.008,0.247,0.300,0.296 |

Table S3. Standardized Mediation Model Results with ADHD as an Outcome

| Predictors | Outcome |  |  |
| --- | --- | --- | --- |
|  | SST Activation in *CNR1* Regions |  |  |
|  | β | SE | p-value |
| Baseline Cannabis Use | -0.350 | 0.661 | 0.597 |
| Sex | 0.071 | 0.08 | 0.371 |
|  | Total ADHD Symptoms |  |  |
|  | β | SE | p-value |
| SST Activation in *CNR1* Regions | -0.286 | 0.126 | 0.023 |
| Baseline Cannabis Use | -0.01 | 0.222 | 0.966 |
| Sex | 0.035 | 0.049 | 0.472 |

Figure S1.
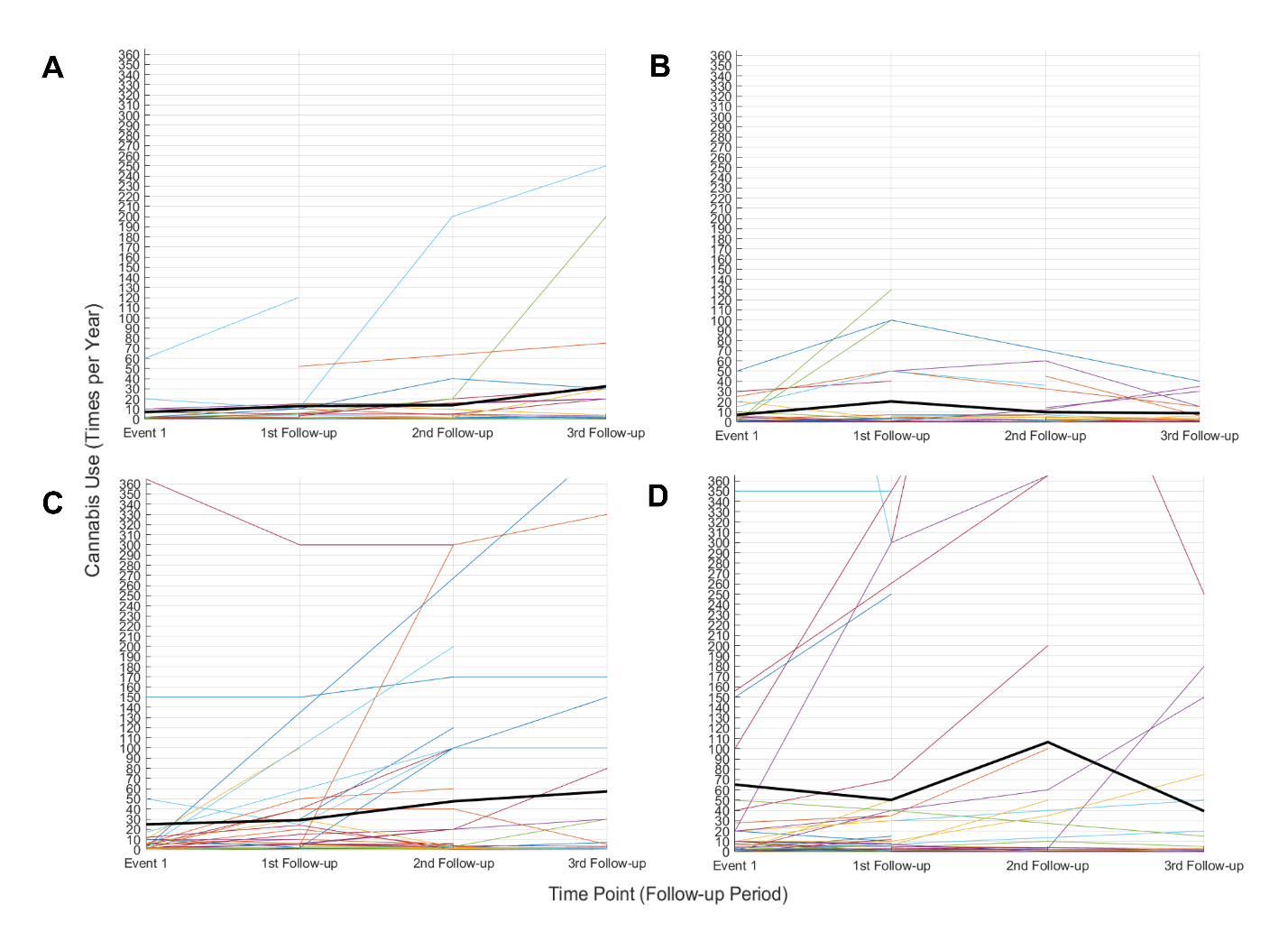

Figure S2.

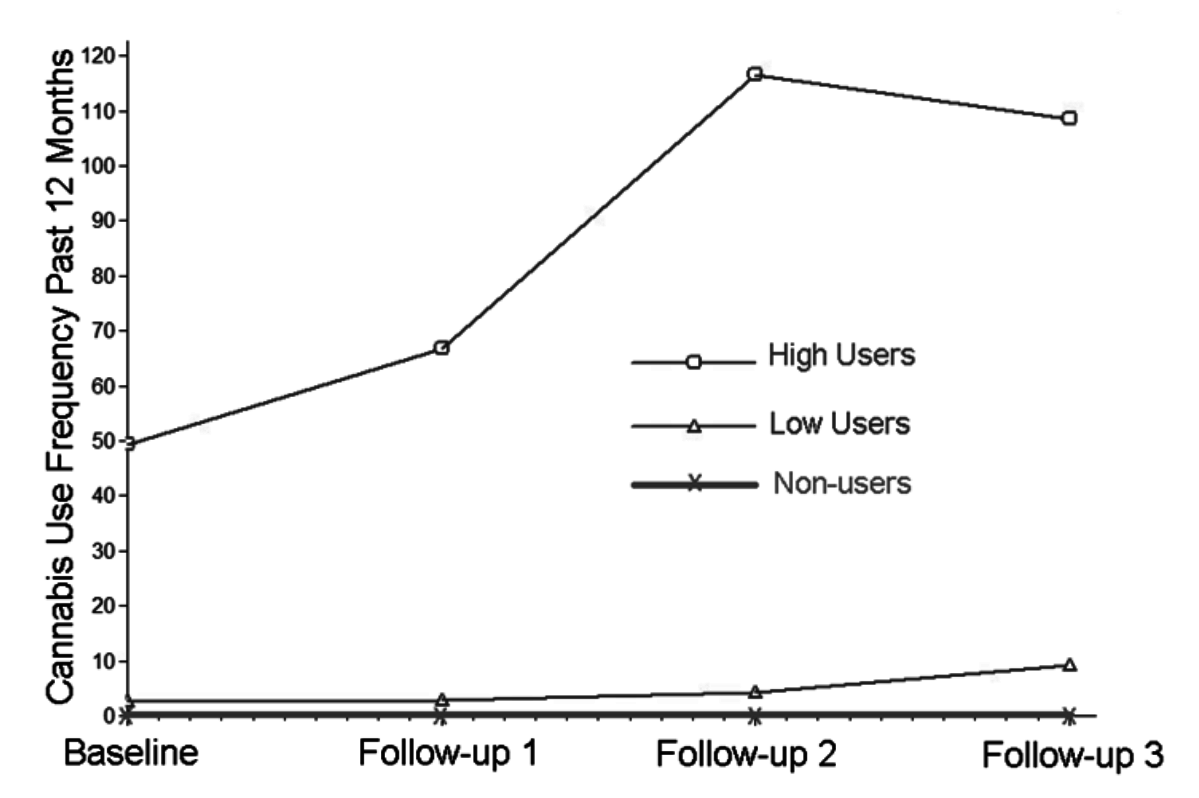

Figure S3.

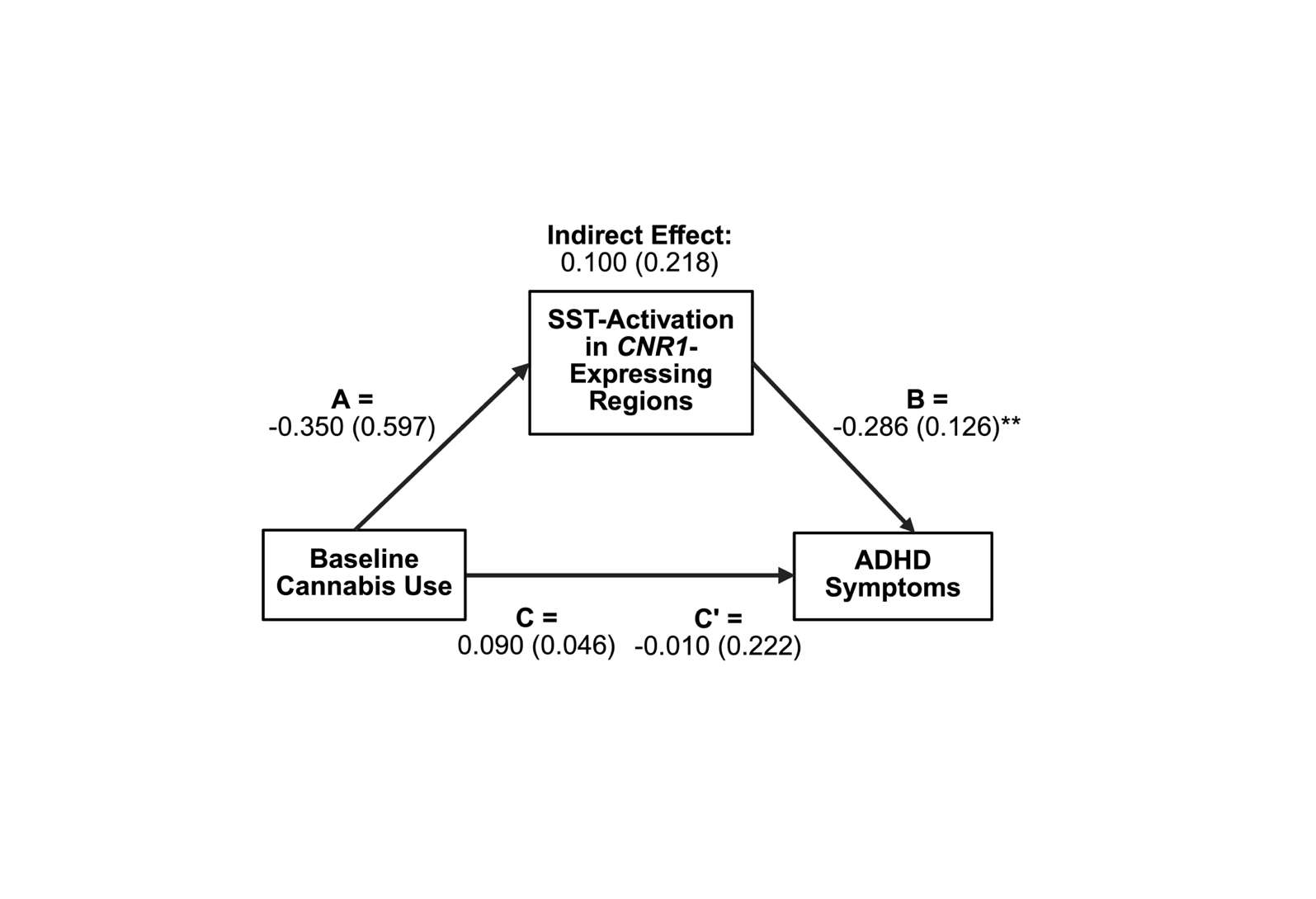

Figure S4.

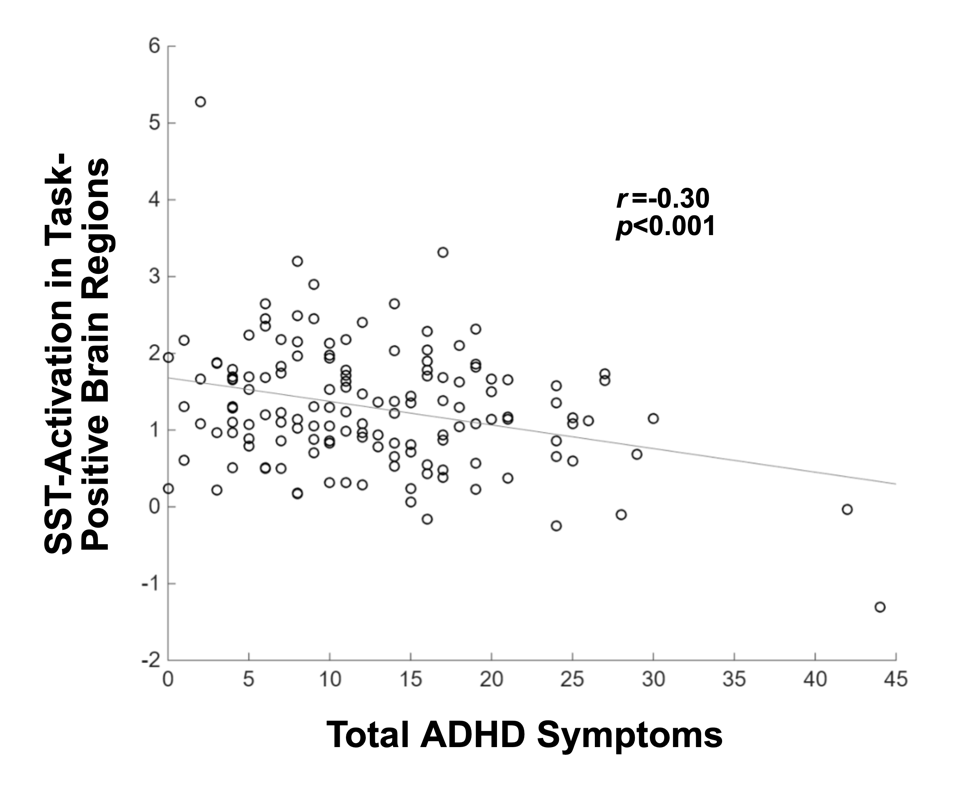

**Supplemental Legends**

Table S1. Growth mixture model class fit statistics. Five latent class models were run to determine the best fit latent class enumeration. Class proportions refer to the sample’s most likely latent class membership for each model.

Table S2: Stop Signal Task Performance descriptive statistics. Mean and standard deviation for calculated performance indices in the neuroimaging sample.

Table S3. Standardized model mediation results with ADHD as an outcome variable. Baseline cannabis use is defined as the self-reported number of times used 1 year prior to the baseline study visit. Total ADHD symptoms are defined by the DSM-IV Total ADHD Symptoms subscale of the CAARS questionnaire. SST-activation in *CNR1* regions is quantified from the covariance of participant SST contrasts and *CNR1*-expression maps.

Figure S1. Longitudinal time series of cannabis use in times per year across each study timepoint. A) Cannabis use in participants in the neuroimaging sample with a CAARS DSM-IV Total ADHD Symptom score below the sample median (≤11). Each line represents a participant with the overlayed bold line showing average cannabis use overtime. B) Cannabis use in participants in the neuroimaging sample with a CAARS DSM-IV Total ADHD Symptom score above the sample median (12-44). C) Cannabis use in participants in the longitudinal sample with a CAARS DSM-IV Total ADHD Symptom score below the sample median (≤15). D) Cannabis use in participants in the longitudinal sample with a CAARS DSM-IV Total ADHD Symptom score below the sample median (16-44).

Figure S2. Growth mixture model latent classes. Model fit statistics led to the selection of a 2-class model composed of high and low cannabis users, with non-users added as a third class. Average cannabis use is plotted over time for each class of cannabis users.

Figure S3. Model of baseline cannabis use mediation of ADHD symptoms through neural activation in *CNR1*-expressing brain regions. Baseline cannabis use does not significantly relate to ADHD symptoms through task activation in regions overlapping with *CNR1-*expression. The results reinforce the prediction that ADHD symptoms predict cannabis use, in contrast to the depicted direction. **p*<0.10, ***p*<0.05, ****p*<0.005.

Figure S4. Scatterplot of mean task-related activation and total ADHD symptoms in task-positive brain regions (*r*=-0.30, *p*<0.001).
